# Transparency in the secondary use of health data: Assessing the status quo of guidance and best practices

**DOI:** 10.1101/2024.08.11.24311808

**Authors:** Olmo R. van den Akker, Robert T. Thibault, John P. A. Ioannidis, Susanne G. Schorr, Daniel Strech

## Abstract

We evaluated what guidance exists in the literature to improve the transparency of studies that make secondary use of health data. To find relevant literature, we searched PubMed and Google Scholar and drafted a list of health organizations based on our personal expertise. We quantitatively and qualitatively coded different types of research transparency: registration, methods reporting, results reporting, data sharing, and code sharing. We found 54 documents that provide recommendations to improve the transparency of studies making secondary use of health data, mainly in relation to study registration (n = 27) and methods reporting (n = 39). Only three documents made recommendations on data sharing or code sharing. Recommendations for study registration and methods reporting mainly came in the form of structured documents like registration templates and reporting guidelines. Aside from the recommendations aimed directly at researchers, we found 31 recommendations aimed at the wider research community, typically on how to improve research infrastructure. Limitations or challenges of improving transparency were rarely mentioned, highlighting the need for more nuance in providing transparency guidance for studies that make secondary use of health data.

## Introduction

Health data has become increasingly accessible to researchers with the advent of large databases providing routine patient data from electronic health records (e.g., OpenSafely, OpenPrescribing, Clinical Practice Research Datalink (CPRD), German Portal for Medical Research Data (FDPG)). The secondary use of health data (SU/HD) for research purposes may yield valuable knowledge, sometimes high risk of bias or even fraud (Mehra, Desai, Kuy, Henry, & Patel, 2020; Mehra, Desai, Ruschitzka, & Patel, 2020; Patel, Desai, Grainger, & Mehra, 2020; Rubin, 2020; Piller & Servick, 2020; The Lancet Editors, 2020) may arise. Because the datasets are not tailor-made to research studies, researchers typically need to inspect the data before being able to develop a sensible analysis plan. However, inspecting the data provides the researchers with information about the variables of interest, thereby potentially biasing the statistical analyses (Orsini et al., 2020). Aside from that, routinely collected health data may be more prone than clinical trial data to selection bias because proper randomization cannot typically be achieved (Beesley & Mukherjee, 2022; Kundu, Shi, Morrison, & Mukherjee, 2023), and to measurement error because of differences in the data entry and classification procedures among health organizations (Young, Conover, & Jonsson Funk, 2018). Furthermore, it can be challenging to identify all available SU/HD studies relevant to a given research question, making it difficult to properly review and synthesize the literature.

The analytical complexity and the potential for bias in SU/HD studies highlights the need for more transparency. A higher level of transparency would allow the scientific community to (1) more easily identify SU/HD studies and thus reduce bias in the review and synthesis of such studies, and (2) more easily identify and correct biases in individual SU/HD studies. Moreover, as secondary use of patient data increasingly works with broad consent or opt-out models, transparency about the studies conducted also plays an important role in building and maintaining social trust in this form of patient data use (Zenker et al., 2022). In a broad consent and opt-out model, patients no longer consent to the individual secondary use studies with their patient data but are only informed about study-wide objectives, risks, and governance of secondary use. In these situations, it is even more important that society is informed about which studies are being conducted.

Important pillars of research transparency are registration, methods and results reporting, and data and code sharing (Westmore et al., 2023). Registration (also called preregistration, see Nosek, Ebersole, DeHaven, & Mellor, 2018; Rice & Moher, 2019) refers to the documentation of research plans (e.g., hypotheses and/or analyses) before research outcomes are known (Hardwicke & Wagenmakers, 2023). Registration allows readers of a scientific paper to assess what the research plan was and whether the author(s) conducted a study as planned. This documentation can help identify potentially biased, data-driven decisions the authors might have made during or after running the study. Moreover, registration of the existence of specific studies has the advantage of making transparent what studies are out there, and thus informing the public, informing researchers working on similar topics, and potentially preventing publication bias (Dickersin & Rennie, 2003). A recent study in the Swedish context shows that only 0.5% of SU/HD studies are prospectively registered (Axfors et al., 2024).

Methods reporting refers to the public documentation of the research goals (e.g., hypotheses) and methods of a scientific study once it is completed. Transparent methods reporting allows readers of a paper to assess whether the study was carried out in line with the registration (assuming a registration is available and sufficiently clear), and potentially rerun and verify analyses or perform other replications (Errington et al., 2021; Ioannidis, 2012; Nosek & Errington, 2020). In the case of SU/HD studies, methods reporting typically involves a detailed description of the handling of data and the statistical analyses performed.

Results reporting refers to the documentation of the outcomes of a scientific study. It is transparent if a result is reported for all the planned analyses, and unplanned analyses are presented as unplanned. Results transparency is important because omitting certain results (e.g., because they are not statistically significant) biases the scientific literature (Hart, Lundh, & Bero, 2012; Kirkham et al., 2010).

Sharing refers to the distribution of the data and code of a study, which can be ‘open’ or ‘controlled’. Open data and open code are available to anyone with access to the internet. Controlled data and controlled code are available to *bona fide* researchers but come with restrictions such as a confidentiality agreement. Controlled sharing is customary for data that are sensitive, which typically is the case for electronic health data. In general, sharing is transparent if it allows readers to redo the study’s analyses on the original data. Data and code sharing are seen as some of the most important transparency practices in biomedicine (Cobey et al., 2023). In the context of SU/HD, control over the data typically lies with the registry or database that provides the data, not with the researchers themselves. Transparency therefore does not necessarily mean providing access to the data but means providing information *how* to access the data from the data provider (if access is possible at all). For example, researchers could provide information about the specifics of the data use agreement they had in place with the data provider, the use-and-access criteria applied by the patient registry, or an explanation why access to the data to third parties is not feasible or allowed. Note that sharing is sometimes called on-sharing in the context of secondary use of data because, per definition, the data has already been shared before (Then et al., 2021).

The importance of research transparency is already well acknowledged in the realm of clinical trials (!Borysowski, Wnukiewicz-Kozłowska, & Górski, 2020), as is evidenced by the large collection of authoritative guidance documents regarding registration (e.g., Gamble et al., 2017; article III.L.1 of International Committee of Medical Journal Editors (ICMJE), 2022; article 35 of World Medical Association, 2013), reporting (e.g., ICMJE, 2022; Schulz, Altman, & Moher, 2010; World Health Organization, 2017; article 36 of World Medical Association, 2013), and sharing (e.g., article III.L.2 of ICMJE, 2022), as well as legal requirements and infrastructure such as clinical trial registries (e.g., https://clinicaltrials.gov, https://www.clinicaltrialsregister.eu)

While there is a large amount of guidance and infrastructure available from international organizations, journals, funders, and research institutions to improve the transparency of clinical trials, the guidance and infrastructure in the area of SU/HD seems less developed. In recent years, some important repositories have taken root that aim to provide health data to researchers (e.g., OpenSafely, Clinical Research Practice Datalink, European Health Data Space). However, to our knowledge, there has been no assessment of peer-reviewed literature or institutional documents regarding guidance for improving the transparency of studies using such data. The current review aims to perform such an assessment.

## Box 1 – Terminology used in this study

### Health data

According to the European Data Protection Supervisor (2023) health data refers to personal information that relates to the health status of a person and includes medical data as well as administrative and financial information about health. Health data can stem from routine clinical processes as well as from patient-reported outcome (PRO) measures (Hjollund, Valderas, Kyte, & Calvert, 2019).

### Secondary use of health data

Researchers have reported some confusion about what secondary use means (Joint Action Towards the European Health Data Space, 2022). We follow the World Health Organization (WHO, 2022) by defining the secondary use of health data as the processing of health data for purposes other than the initial purposes for which the data were collected. Even though health data can have many secondary uses (Safran et al., 2007) we only focus on its use for biomedical research. A largely synonymous term that has gained traction in recent years is ‘real-world data’. Real-world data is typically used to refer to health data that is not derived from clinical trials but during routine clinical practice (Makady, De Boer, Hillege, Klungel, & Goettsch, 2017).

### Transparency

We use transparency in the context of scientific research, by focusing on registration, reporting, and sharing. However, transparency in the context of SU/HD is often also used to mean transparency with regard to the patient (i.e., whether the patient knows what happens with their personal data, see e.g. Geissbuhler et al., 2013; Hripcsak et al., 2014). The ethical and legal debate on whether patients should be informed about every secondary use project involving their patient data to decide whether to give their consent is not addressed in this paper (but see Zenker et al., 2022).

## Methods

The study design was registered on 20-07-2023 on the Open Science Framework at https://osf.io/7864h. The raw data and analyzed data used in this study can be found at https://osf.io/2nup4. Note that we also preregistered an assessment of the transparency guidance on patient registry websites but in hindsight we realized that providing guidance is not one of the main goals of such websites. As such, we do not present results from that part of the preregistration in this paper. Additionally, we decided not to pursue our preregistered analysis on dataset registration (i.e., publicly registering that and how you are using a specific dataset). We did so because we could not find any references to dataset registration in the first batch of about 30 documents and decided it would not be worth the extra coding effort to further pursue it. An overview of all deviations from our preregistration can be found at https://osf.io/m4ehx.

### Sample selection

#### Peer-reviewed literature

To find documents that potentially discuss transparency in SU/HD studies, we used the following search term combination on PubMed: (“guidance” OR “best practice*” OR “guideline*” OR “recommendation*” OR “road map” OR “position paper”) AND (“secondary use” OR “secondary data” OR “reuse” OR “database stud*” OR “real-world data” OR “real-world evidence” OR “registry data”) AND (“transparen*” OR “registration” OR “reporting” OR “sharing”). This PubMed search retrieved 954 documents (see https://osf.io/6k879) on 20-07-2023. We also did a search of these keyword combinations on Google Scholar, where we added the term “health” to restrict our search to the secondary use of health data.

Prior to registration, we conducted the search process using Google Scholar. Unfortunately, search results for Google Scholar are not reproducible (Gusenbauer & Haddaway, 2020). We decided to include Google Scholar despite the disadvantage of irreproducibility because it is the most comprehensive (Gusenbauer, 2019; Martín-Martín, Thelwall, Orduna-Malea, & López-Cózar, 2021) and most used (Nicholas et al., 2017; Van Noorden, 2014) source of scientific literature. Moreover, Google Scholar is especially useful for exploratory searches like ours (Athukorala, Głowacka, Jacucci, Oulasvirta, & Vreeken, 2016). To identify any missed documents that may be relevant, we used the snowball method and searched the references section of the documents included based on the initial screening.

We also wanted to include documents from health institutions with a relevance for SU/HD studies. Based on our own expertise and the overview provided by Burns et al. (2022) we selected a set of (inter)national health institutions that had previously published guidance on clinical trials, and a set of learned societies specifically revolving around SU/HD studies. We then looked on their websites for any documents that may conceivably include transparency guidance for SU/HD studies.

From all identified documents (both peer-reviewed and institutional), we selected documents relevant to our research question in the following way. First, we screened the title and abstract of a document and assessed whether the document itself was likely to contain guidance for any of the transparency aspects: registration, methods reporting, results reporting, data sharing, and code sharing. We assessed that this would be the case for documents that state:

– that they provide guidance for SU/HD studies on one or more transparency aspects,
– that they provide general guidance for SU/HD studies,
– that they discuss one or more transparency aspects in the context of SU/HD studies, and
– that they discuss SU/HD studies generally.

In sum, our set of included documents involved documents that based on the title and abstract potentially included transparency guidance for SU/HD. The full set of peer-reviewed papers and institutional documents can be found at https://osf.io/ednwx and https://osf.io/gajxt, respectively.

### Analysis

We employed both a qualitative as well as a quantitative approach. The qualitative approach consisted of a thematic analysis (Braun & Clarke, 2006) using MAXQDA (VERBI Software, 2021) in which we retrieved relevant sections from our sample of selected documents. In the first stage of coding, our approach was primarily deductive as we screened documents for text relevant to one of the transparency themes: registration, methods reporting, results reporting, data sharing, and code sharing. We also extracted texts that highlighted the main goal of the documents, and any additional texts that we deemed potentially useful in writing our paper.

In the second, inductive stage of coding, we identified subthemes within the *a priori* selected themes. We went over all the extracted texts from a given theme and categorized each text based on content. This led to a more granular understanding of guidance in the main transparency areas.

The quantitative approach consisted of counting the number of documents that included one or more texts regarding each of the themes we used in the deductive stage, and all the subthemes identified in the inductive stage of our thematic analysis. We distinguished three ways in which the themes were included in a document. First, we noted when the authors made a general call related to any of the themes or sub-themes. We defined a general call as a statement that the transparency in a certain area should be improved, but without providing a rationale for this statement. Second, we noted when the authors justified the need for improved transparency in a certain area. Third, we noted when the authors made a recommendation for *how* to improve the transparency in a certain area. If a document contained multiple calls for a particular transparency practice, we coded this as one call because calls do not qualitatively differ from one another like justifications and recommendations do (e.g., there could be multiple justifications for why we need more registration of SU/SH studies). As such, a document could have a maximum of five calls, one for each of the transparency practices, but in theory unlimited justifications and recommendations.

In some cases, we initially coded a text excerpt as a general call because authors did not directly explain why more transparency would be beneficial. However, justifications were often provided earlier or later in the text. In those cases, we recoded the ‘call’ to a ‘justification’. Consequently, our dataset does not include documents with a call *and* a justification. Do note that a justification does not automatically imply a call, and a recommendation does not automatically imply a justification or a call. For example, it could be that a paper states that a health organization has called for more registration and then provides recommendations, which does not mean that the authors in the paper call for more registration.

## Results

Below we present the quantitative and qualitative results in narrative form. In addition, we present the quantitative results in tabular form in Table 1, and visually in Figure 1 (peer-reviewed literature) and Figure 2 (institutional documents), and the qualitative results in tabular form in Table 2 (registration), Table 3 (reporting), and Table 4 (sharing), and visually in Figure 3 (justifications) and Figure 4 (recommendations).

**Figure 1.**
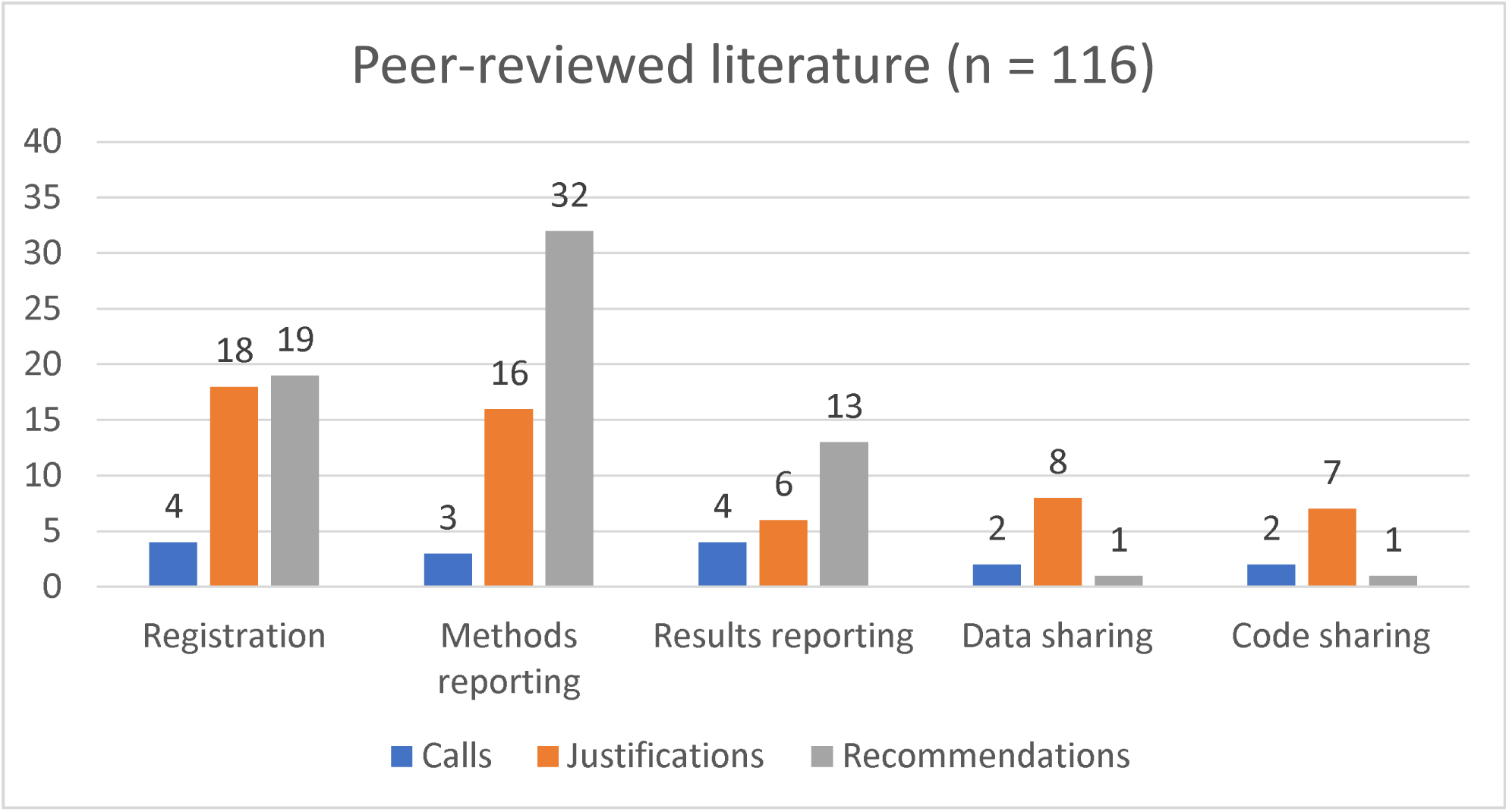
Number of Papers with at least one Call, Justification, or Recommendation regarding the Five Transparency Elements in the Peer-Reviewed Literature.

**Figure 2.**
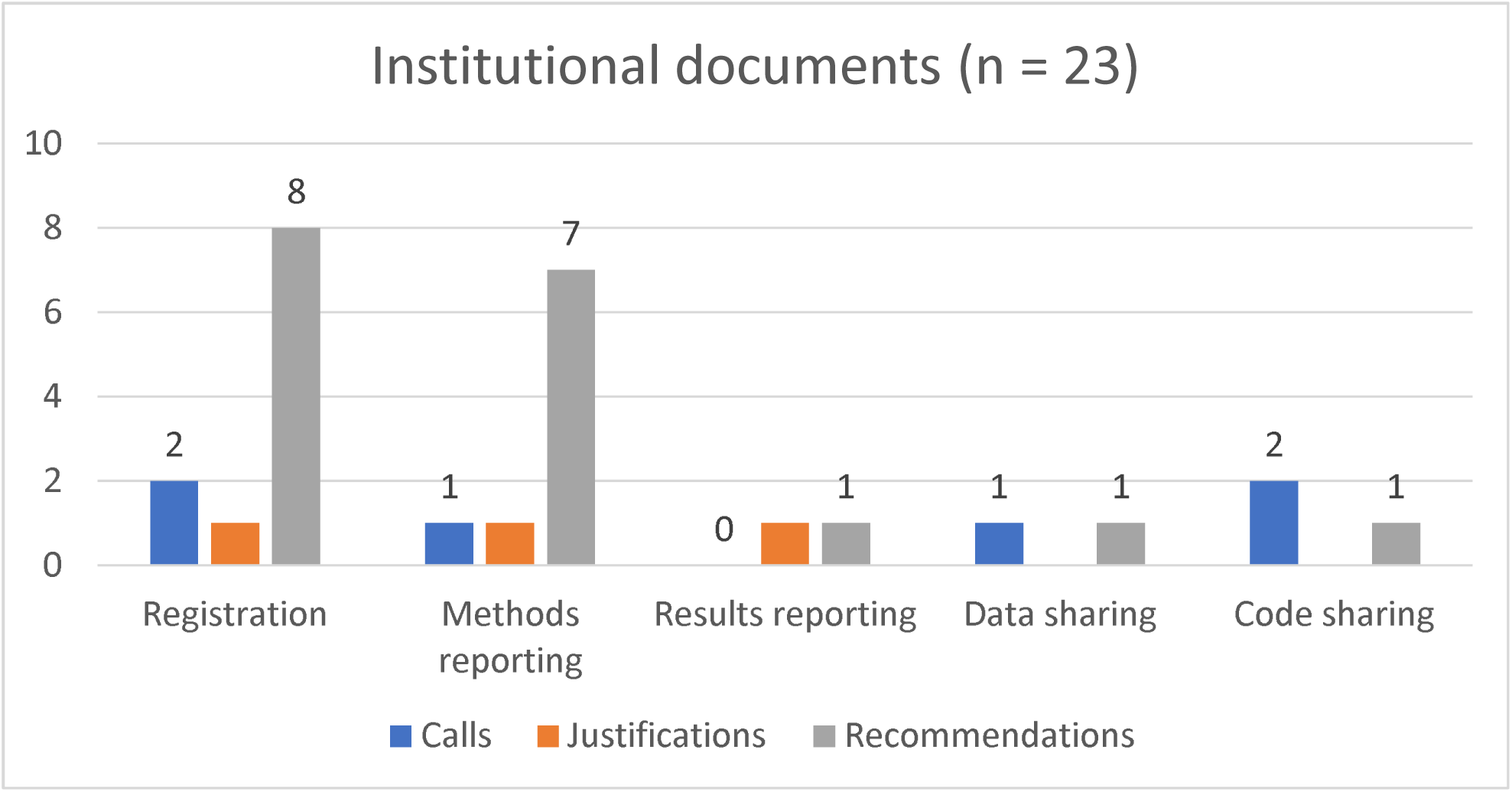
Number of Documents with at least one Call, Justification, or Recommendation regarding the Five Transparency Elements in Institutional Documents.

**Figure 3.**
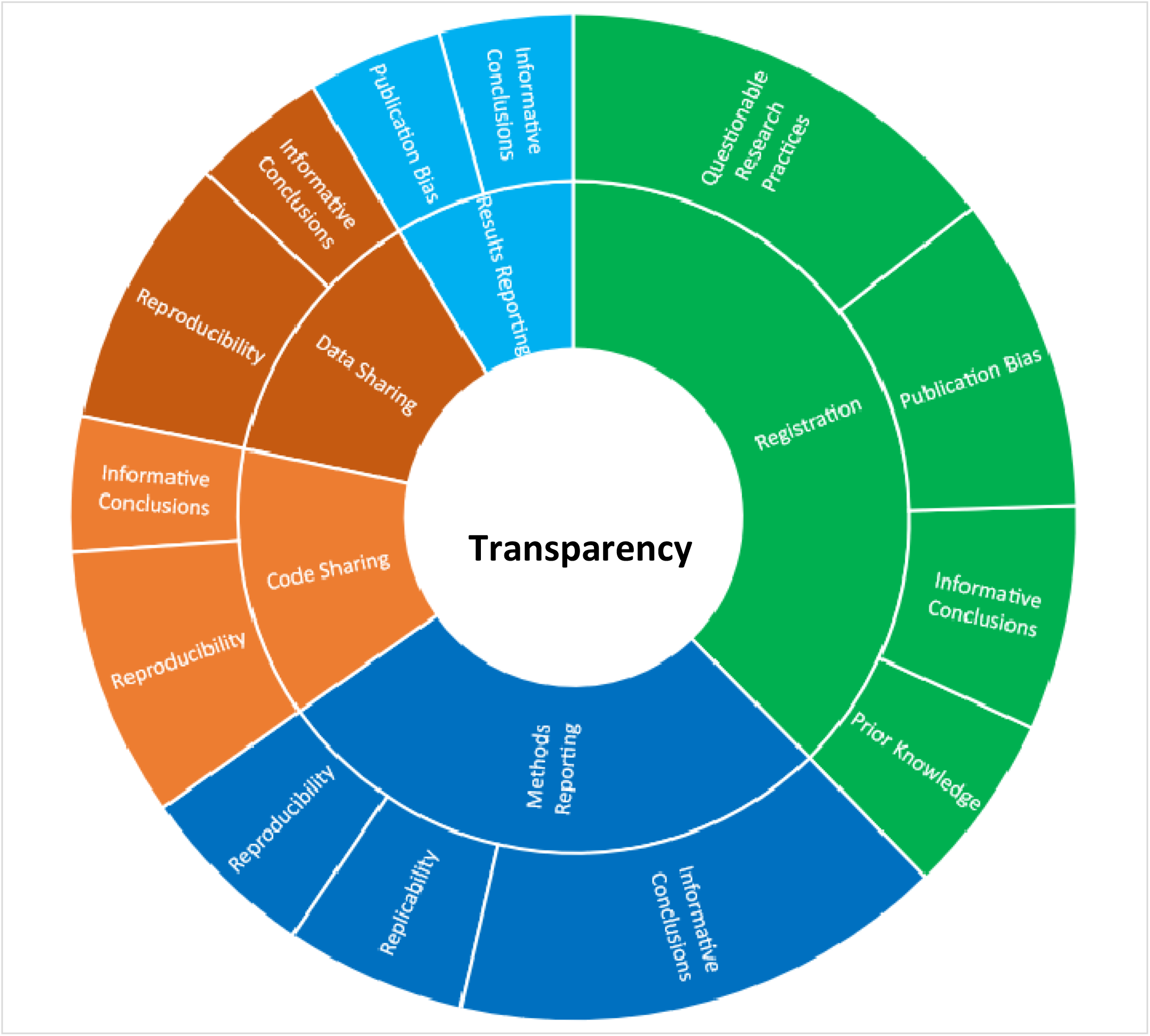
Overview of the Justifications in Peer-Reviewed Literature and Institutional Documents for Improving the Transparency in Studies Making Secondary Use of Health Data *Note*. The sizes of the areas represent the number of times we found these justifications in the literature

**Figure 4.**
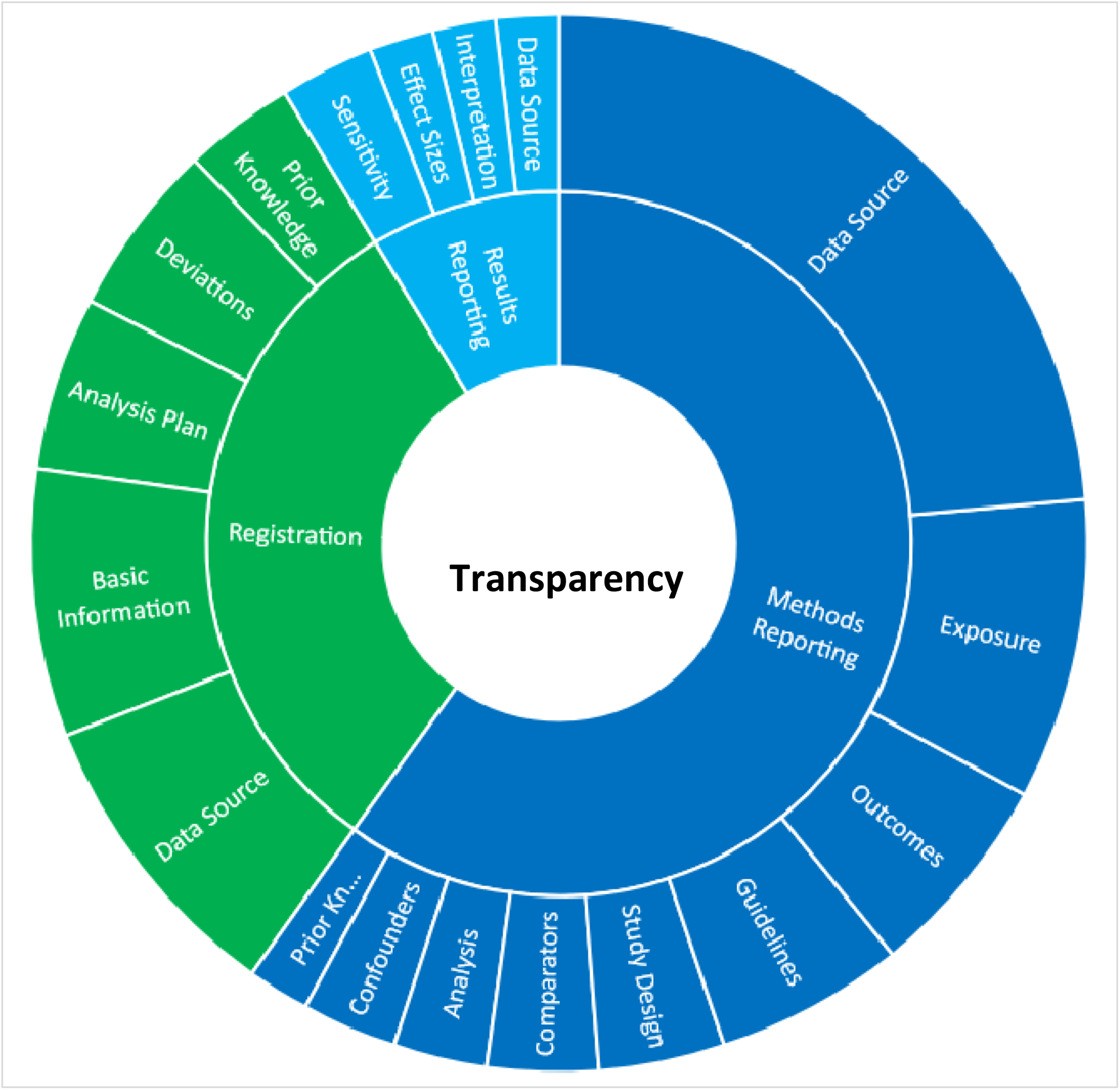
Overview of the Recommendations in Peer-Reviewed Literature and Institutional Documents for Improving the Transparency in Studies Making Secondary Use of Health Data *Note*. The sizes of the areas represent the number of times we found these recommendations in the literature

**Table 1.** Total Number of Calls (C), Justifications (J), and Recommendations (R) in the Peer-Reviewed Literature and Institutional Documents.

|  | Peer-Reviewed Literature<br>(n = 116) |  |  | Institutional Documents<br>(n = 23) |  |  |
| --- | --- | --- | --- | --- | --- | --- |
|  | C | J | R | C | J | R |
| Registration | 4 | 24 | 112 | 2 | 1 | 55 |
| Methods Reporting | 3 | 24 | 147 | 1 | 1 | 19 |
| Results Reporting | 4 | 6 | 26 | 0 | 1 | 1 |
| Data Sharing | 2 | 9 | 1 | 1 | 0 | 1 |
| Code Sharing | 2 | 9 | 4 | 2 | 0 | 1 |
| Total | 15 | 72 | 290 | 6 | 3 | 77 |

**Table 2.** Qualitative Analysis of Justifications and Recommendations on Registration.

|  | Explanation | Example quote |
| --- | --- | --- |
| **Justifications** |  |  |
| Publication Bias | Registration can prevent or at least allows identification of publication bias because it provides a record of studies that are not published | “How can investigators presenting an RWE study dispel the suspicion that they ran multiple similar studies and analyses, but published only the one that gave a positive result? One answer is to adopt institutional and corporate policies on RWE studies that provide a similar level of rigor to policies on the conduct of RCTs. Such a policy may set the definition of RWE studies, mandate posting an outline protocol for each study on an appropriate forum.” (White, 2017) |
| Questionable Research Practices | Registration can prevent or at least allows identification of questionable research practices because registration allows third parties to compare the plans to the actual study | “Because research plans and hypotheses are specified before the results are known, pre-registration reduces the potential for cognitive biases to lead to p-hacking, selective reporting, and HARK-ing.” (Baldwin et al., 2022) |
| Informative Conclusions | Registration allows third parties to better assess the study and its results when the study has been completed | “[T]ransparency improves the ability of decision-makers to assess the quality and validity of a study by giving them a deeper understanding of why and how the research was conducted and whether the results reflect pre-established questions and methods.” (Orsini et al., 2020) |
| Prior Knowledge | Registration makes clear what prior knowledge authors had about the dataset that could have biased their decisions | “Bias resulting from retrospective selection can be serious, especially when selecting external data and key analysis features, when the external control results are already known. Pre-specification is therefore an essential pre-requisite when using external control data” (Burger et al., 2021) |
| **Recommendations** |  |  |
| Deviations | Transparently discuss any deviations made from the registration in the final paper | “We recommend that researchers be transparent about their ex ante analytic plans, provide justification for subsequent changes in analytic models, and report out the results of their ex ante analytic plan as well as the results from its modifications.” (Berger et al., 2009) |
| What To Register: |  |  |
| - Basic Information | Provide basic information about the study, like the study rationale and the main hypotheses | “Please provide the hypotheses of your secondary data analysis. Make sure they are specific and testable, and make it clear what your statistical framework is (e.g., Bayesian inference, NHST). In case your hypothesis is directional, do not forget to state the direction. Please also provide a rationale for each hypothesis.” (Van den Akker et al., 2021) |
| - Data Source | Provide information about the data source / study population | “First, registration would require researchers to prespecify their data source(s) along with sample inclusion and exclusion criteria allowing preanalytic evaluation of whether the study is representative of the patient population using the medical product. For any data source, quality-control measures to ensure data integrity would be proactively described.” (Dhruva et al., 2020) |
| - Prior Knowledge | Provide information about any knowledge the authors have about the data | “To increase transparency about potential biases arising from knowledge of the data, researchers could routinely report all prior data access in a pre-registration. This would ideally include evidence from an independent gatekeeper (e.g., a data guardian of the study) stating whether data and relevant variables were accessed by each co-author.” (Baldwin et al., 2022) |
| - Analysis Plan | Provide detailed information about the planned statistical analyses | “Describe details of sensitivity analysis that will be performed to confirm the robustness of the results of analysis. Sensitivity analysis is especially important in pharmacoepidemiological studies with databases because the results of analysis tend to vary significantly depending on study design such as definition of exposure, outcome, covariates, etc. Describe all previously planned sensitivity analyses and ensure that these are differentiated from additional interim sensitivity analyses.” (Japanese Pharmaceuticals and Medical Devices Agency, 2014) |

**Table 3.** Qualitative Analysis of Justifications and Recommendations on Reporting.

|  | **Explanation** | **Example** |
| --- | --- | --- |
| **Methods Reporting** |  |  |
| ***Justifications*** |  |  |
| Informative Conclusions | Methods reporting provides context with which third parties can interpret the drawn conclusions in a less biased manner | “Data users should agree to make the methods and results of their secondary analyses publicly available not only through scientific publications (that may or may not be prepared and, if prepared, that may or may not be accepted for publication) but also by depositing them in a repository and making them discoverable. This will be important to provide further examples of effective data sharing and allow any conclusions from secondary use to be examined by others.” (Ohrmann et al., 2017) |
| Reproducibility | Providing information about the methods / statistical analyses helps other researchers to redo the analysis using the same dataset to see whether the results are consistent | “Because of the lack of standardization in secondary data analytics, complete transparency is critically important in the reporting of analytic approaches and all coding details. This will allow reproduction of analyses, replication of findings using different data sources, and ultimately greater confidence in such analyses, possibly approaching the trust we place in highly controlled clinical trials.” (Schneeweiss, 2016) |
| Replicability | Providing information about the methods / statistical analyses helps other researchers to redo the analysis using a different dataset to see whether the results are consistent | “This system would enable regulators to repeat the exact same study and change assumptions or definitions in the design and statistical analysis either through submission of data or by providing access to the data.” (Franklin et al., 2019) |
| ***Recommendations*** |  |  |
| Guidelines | Use existing reporting guidelines to write the methods section of your research papers | “Several guidelines have been developed to enhance reporting, such as Strengthening the Reporting of Observational Studies in Epidemiology (STROBE), the Reporting of studies Conducted using Observational Routinely-collected Data (RECORD) statement, and its extension for pharmacoepidemiology studies (RECORD-PE). Interested researchers should always consult these |
|  |  | guidelines for reporting of their studies.” (Liu et al., 2021) |
| **What To Report:** |  |  |
| - Overview study design | Provide a descriptive or visual overview of the basic design elements that make up your study | “Reporting on overall study design should include a figure that contains 1<sup>st</sup> and 2<sup>nd</sup> order temporal anchors and depicts their relation to each other.” (Wang et al., 2017) |
| - Data Source / Study Population | Provide any relevant details about where the data came from and how they were managed | Describe the nature of dataset(s) used. In particular: The purpose of the dataset – e.g. observational research registry, national audit programme, administrative dataset (linked to financial remuneration or service delivery). This should include details of the funding of the dataset, and the organisation(s) responsible for the administration and oversight.” (Perry et al., 2014) |
| - Prior Knowledge | Provide any prior knowledge about the data that the authors may have had before doing the statistical analyses | “Authors should describe the extent to which the investigators had access to the database population used to create the study population.” (Benchimol et al., 2015) |
| - Exposures / Predictors | Provide information about the factor, condition, or intervention that may impact the health outcomes of interest | “Reporting on exposure definition should include: The type of exposure that is captured or measured, e.g. drug versus procedure, new use, incident, prevalent, cumulative, time-varying.” (Wang et al., 2017) |
| - Comparators / Control Sample | Provide information about the groups or conditions against which the exposure group is compared to evaluate the impact of the exposure on the health outcome of interest. | "When compared with another TCM exposure / intervention, it is necessary to evaluate whether the evidence base for the efficacy and safety of the control is sufficient." (Chai et al., 2022) |
| - Confounders / Covariates | Provide information about the variables that are (potentially) associated with both the exposure and the health outcome of interest | “Discuss the potential for confounding, both measured and unmeasured, and how this was assessed and addressed.” (Berger et al., 2009) |
| - Outcomes | Provide information about health-related endpoints or events of interest | “Discuss how outcomes were measured and how classification bias was addressed.” (Berger et al., 2009) |
| - Statistical Analysis | Provide detailed information about the statistical methods used to draw inferences about the variables of interest | "Describe the methods used to evaluate whether the assumptions have been met." (Langan et al., 2019) |
| **Results Reporting** |  |  |
| **Justifications** |  |  |
| Informative Conclusions | Effective reporting of results helps make more | “The results of all analyses that are conducted (e.g., |
|  | accurate conclusions of the results of a study | matched, unmatched, adjusted, and unadjusted) results should be reported. Presentation of the unadjusted results helps to demonstrate the robustness of the chosen method of analysis; matched or adjusted results that differ substantially from the unmatched/unadjusted can reduce confidence in the matched/adjusted trends observed.” (Rochet et al., 2013) |
| Publication Bias | Also presenting non-statistically significant results helps to prevent the literature from being disproportionally filled with “positive” studies | “The lack of RW study protocol registration and reporting of results can potentially lead to significant bias in reporting positive/selective results, as studies that do not produce the expected data will probably not be completed or submitted for peer review. RW studies, regardless of the origin of RW data, need to be registered in a manner equivalent to that of clinical trials.” (Van den Broek et al., 2022) |
| **Recommendations** |  |  |
| Patient Characteristics | Provide detailed information about the characteristics of the patients in the sample | “The rationale for reporting on characteristics of the study population is described in numerous other reporting guidance documents. This includes items such as an attrition table (showing patient numbers as eligibility criteria are applied), baseline characteristics of the derived population, as well as the number and timing of outcomes of interest. It allows the investigator and reviewers to describe and assess whether the frequency of a derived variable is consistent with expectation (e.g., that the outcome incidence or a covariate prevalence looks approximately correct). The same rationale applies in studies that develop or use derived information from NLP and ML algorithms.” (Wang et al., 2019) |
| Sensitivity Analyses | Provide more than one analysis so that others can assess the robustness of the results | Report univariate and multivariate results in an unbiased and complete fashion such that the benefits and risks of all comparators reflect “fair balance.” (Willke & Mullins, 2011) |
| Effect sizes | Provide effect sizes alongside the statistical significance of the results | “Second, we recommend researchers report effect sizes as many associations may be statistically, but not practically, significant when analyzing large sample sizes. In doing so, |
|  |  | we may need to adjust our collective expectations of what effect sizes to expect, and which ones to treat as substantial” (Kievit et al., 2022) |
| Interpretation | Provide a cautious discussion of the results in light of the research questions and/or hypotheses. | “Discuss the potential for confounding by indication, contraindication or disease severity or selection bias (healthy adherer/sick stopper) as alternative explanations for the study findings when relevant.” (Langan et al., 2018) |

**Table 4.** Qualitative Analysis of Justifications and Recommendations on Sharing.

|  | Explanation | Example quote |
| --- | --- | --- |
| **Data sharing** |  |  |
| *Justifications* |  |  |
| Informative Conclusions | Shared data, including metadata, allows third parties to make better assessments of the findings based on the data | “The disclosure of research-related information is also fundamental to improving the transparency of studies, especially the availability of raw data, which enables readers to assess the authenticity and reliability of the findings.” (Zhao et al., 2023) |
| Computational Reproducibility | Data sharing (and code sharing) allows third parties to redo the analyses in the study to see whether the same results are found | “For full analytic reproducibility, sharing of code and data is encouraged. However, there are often privacy and intellectual property considerations that prevent sharing of data, data derivatives, or code.” (Wang et al., 2019) |
| Sensitivity Analyses | Data sharing allows third parties to do analyses that differ from the analyses in the original study, allowing an assessment of the reliability of the results | “Ideally reviewers of submitted evidence, including HTA bodies or independent review groups, would also have access to the data and analytical code to ensure the replicability of the submitted results and assess the impact of alternative analytical decisions or data on the resulting estimate(s). However, there remain substantial governance, technical and practical challenges to sharing data, including a lack of in-house expertise in many HTA agencies.” (Kent et al., 2021) |
| **Code sharing** |  |  |
| *Justifications* |  |  |
| Informative Conclusions | Shared code allows third parties to see the exact analyses that were conducted, allowing a better | “Finally, the analyses conducted on secondary data are commonly more complex than those applied to simpler |
|  | interpretation of the results | experimental designs. Methods sections in high-impact journals are often highly condensed or hidden at the end of an article, which can make it difficult or even impossible to assess which analyses exactly were performed. To address this issue, we recommend authors always publish the full analytic code, even when the raw data cannot be directly shared.” (Kievit et al., 2022) |
| Computational Reproducibility | Shared code allows third parties to redo the analyses in the study to see whether the same results are found | “Irreproducibility can be mitigated by sharing raw and processed data and codes, assuming no privacy is compromised in this process. For replicability, given that RWD are not generated from controlled trials and every data set may has its own unique data characteristics, complete replicability can be difficult or even infeasible. Nevertheless, detailed documentation of data characteristics and pre-processing, pre-registration of analysis procedures, and adherence to open science principles (e.g., code repositories) are critical for replicating findings on different RWD datasets, assuming they come from the same underlying population.” (Liu & Demosthenes, 2022) |

*Note.* In contrast to Table 2 and Table 3, this table does not include recommendations because we could not find enough recommendations to do a thematic analysis.

### Guidance in the peer-reviewed literature

Our sample of peer-reviewed literature (which was slightly different from our preregistered sample, see https://osf.io/m4ehx) that included guidance on transparency for SU/HD studies contained 116 papers (36 first found through Google Scholar, 57 first found through PubMed, and 23 found through the snowball method by checking the 4,745 references of the papers found through Google Scholar and PubMed). We extracted 606 text excerpts from the peer-reviewed papers. The prevalence of calls, justifications, and recommendations in those excerpts is provided in Table 1 and in the sections below.

#### Registration

Among the 116 papers, we found four papers with a call for more registrations of SU/HD studies, eighteen with a justification for more registrations of SU/HD studies (of which four had multiple justifications), and nineteen with one or more recommendations on how to register SU/HD studies. In total, we found 112 recommendations, with three papers providing most (61/112) of those (25 in Van den Akker et al., 2021; twelve in Wang et al., 2021; and 24 in Wang et al., 2022). These three papers involved structured templates that researchers can use to register SU/HD studies. We counted each individual item of these templates as a separate recommendation, but we excluded subitems. For example, for Wang et al. (2017) we coded the reporting item “Methods used for confounder adjustment” as a separate recommendation but did not code the subitems in which the method for confounder adjustment was discussed for different potential statistical analyses.

##### Justifications

The main justification for registering SU/HD studies was that registration would prevent questionable research practices like *p*-hacking, selective or retrospective reporting, and HARKing (Hypothesizing After the Results are Known). Several authors argued that the large scale (Van den Akker et al., 2019; Zarin et al., 2020) and widespread availability of health datasets (Orsini et al., 2020; Van den Akker et al., 2019) make analyses based on existing health data more susceptible to biases because of the large number of analysis options and possible prior knowledge of the data. Another commonly mentioned justification for registration was that it would allow someone to identify publication bias or potentially prevent publication bias. This is because an overview of all published and non-published studies allows the research community to identify non-published studies and request the results to be made available. The authors of one study claimed that publication bias might be more severe for SU/HD studies because “*journals may have less expertise in evaluating such studies*” (Orsini et al., 2020). Finally, a paper mentioned that registration could be helpful in drafting ethical review board submissions and informed consent forms (Santos et al., 2017) and justified more registrations of SU/SH studies on that ground.

##### Recommendations for Researchers

Most often, recommendations on how to register SU/HD were about which study elements need to be specified in a registration. Many elements were mentioned but in the context of SU/HD most emphasis was placed on registering the data source and the statistical choices. Registering the authors’ prior knowledge of the data was emphasized strongly by Baldwin et al. (2022), Orsini et al., (2020), and Van den Akker et al. (2021), but was not mentioned in two extensive papers presenting registration guidance by Wang et al. (2021; 2022). Several papers simply provided a list of platforms where authors can preregister their SU/HD study, where references to clinicaltrials.gov and the electronic Register of Post-Authorization Studies were most common (Berger et al., 2017; Moon et al., 2023; Orsini et al, 2020; Santos et al., 2017). Finally, several papers stated that deviations from registrations should be transparently disclosed, preferably including the timing of and justification for the change (e.g., Baldwin et al., 2022; Berger et al., 2009; Dhruva et al., 2020; Kent et al., 2021; Orsini et al., 2020; Wang et al., 2022a; Wilke & Mullins, 2011).

##### Recommendations for Institutions

We also found several recommendations in the literature where guidance was provided to institutions on how to improve the infrastructure surrounding registration. For example, Orsini et al. (2020) discussed that embargoing registrations could preserve intellectual property and prevent scooping. Zarin et al. (2020) and Wang et al. (2021) focused more on registration templates, where the former argued for creating balanced templates that take into account both comprehensiveness (providing more information about the study) and simplicity (yielding higher uptake of registration of SU/HD studies), and the latter argued for the integration of templates with HD registries.

##### Limitations

Some authors also provided points of concern with relation to the registration of SU/HD studies, although these discussions were often limited. Orsini et al. (2020) stated that registration does not guarantee high-quality studies, and Zarin et al. (2020) argued that registration of SU/HD studies likely has limited impact unless any recommendations or policies can be legally enforced. Dhruva et al. (2020) agreed with the point about enforcement and called for a mandate for registration of SU/HD studies as the 2004 ICMJE policy did for clinical trials (De Angelis et al., 2004).

#### Reporting

We found three papers with a call for methods reporting, four with a call for results reporting, sixteen with a justification for methods reporting, six with a justification for results reporting, 32 with one or more recommendations for how to best report the methods of a SU/HD study, and thirteen with one or more recommendations for how to best report the results of a SU/HD study. In total, we found 147 recommendations for methods reporting, and 26 recommendations for results reporting. Five papers provided more than ten recommendations (thirteen in Benchimol et al., 2015; fifteen in Chai et al., 2022; thirteen in Langan et al., 2018; 50 in Wang et al., 2017; eleven in Wang et al., 2019).

##### Justifications

Justifications for better methods reporting typically came in two shapes: (1) good reporting provides the information necessary to reproduce (i.e., redo the analyses on the same dataset) or replicate (i.e., do the same analyses on a different dataset) results from SU/HD studies (e.g., Berger et al., 2017, Denaxas et al., 2017; Schneeweiss et al., 2016, Wang et al., 2017), and (2) good reporting makes it easier to assess the results of the study itself by other researchers or peer reviewers (e.g., Ohmann et al., 2017; Perry et al., 2014; Santos et al., 2017, Wang et al., 2023; White, 2017).

Several authors justified results reporting by claiming that it is a good way to provide information to patients, enhancing trust and facilitating informed decisions (e.g., Cave et al., 2019; Cumyn et al., 2023; Van den Broek et al., 2022). Others mentioned that reporting the results of all conducted analyses would decrease publication bias (White, 2017; Van den Broek et al., 2022).

##### Recommendations for Researchers

The main recommendations for improved methods reporting came in the shape of formal reporting guidelines, in which authors presented lists of relevant study elements that are important to include in research reports. Some papers presented a guideline themselves (Benchimol et al., 2015; Langan et al., 2018; Nicholls et al., 2015; Wang et al., 2017), while others merely advised to adhere to such guidelines (Heikinheimo, 2017; Khosla et al., 2018; Liu et al., 2022; Nicholls et al., 2015; White, 2017). The Reporting of Studies Conducted Using Observational Routinely-Collected Health Data, RECORD (Nicholls et al., 2015) and Wang et al. (2017) had a general goal to improve reporting in SU/HD studies. Other guidelines were more specific: RECORD-PE (Langan et al., 2018) is dedicated to improving reporting in pharmacoepidemiologic research, Patorno et al. (2020) discussed RECORD in light of the reporting of diabetes research, and Chai et al (2022) focused on traditional Chinese medicine.

Recommendations for results reporting were limited because many authors simply state *that* results should be reported and *why*, not necessarily *how* (though an elaborate list of different elements to consider can be found in the Supplementary Materials of Wang et al., 2017). Berger et al. (2018) stated that not only medical journals can be used to report results, but also publicly available websites. Hersh et al. (2013), Roche et al. (2013), and Wang et al. (2019) emphasized that sensitivity analyses can be useful when presenting results because the many analysis options in secondary health data give rise to many different interpretations of the data.

##### Recommendations for Institutions

We also found some institutional recommendations regarding reporting. For example, while the value of reporting guidelines was echoed by Khachfe et al. (2021) they emphasized that such guidelines should be included in manuscript submission and editorial processes for them to be effective. In a similar way as Dhruva et al. (2020) did for registration, they stated that the ICMJE could play a mandating role in this regard. In addition, Khachfe et al. (2021) argued that more domain-specific checklists should be drafted, and such checklists could already be integrated in educational modules. Finally, Bate (2017) mentioned that reporting guidelines for unstructured data like social media data are lacking and that meta-research on the impact of the guidelines is desirable.

##### Limitations

A limited number of concerns or critiques were raised. Both Kent et al. (2021) and Orsini et al., (2020) warn that adhering to reporting guidelines does not necessarily support reproduction, and that they are not necessarily a sign of high research quality.

#### Sharing

Guidance on sharing the data and sharing the code of SU/HD studies was sparse in the peer-reviewed literature. Two papers included a call for more data sharing and more code sharing, eight papers included a justification for more data sharing, and seven papers for more or better code sharing. We found one recommendation in a paper with regard to data sharing (Matandika et al., 2020), and four recommendations in one paper (Denaxas et al., 2017) with regard to code sharing.

##### Justifications

As justifications for data and code sharing, we found that it would allow computational reproducibility checks (e.g., Liu & Demosthenes, 2022, Kievit et al., 2022; Wang et al., 2021) and robustness checks (e.g., Franklin et al., 2019; Kent et al., 2021). Both have to do with redoing the analysis but the goal of computational reproducibility checks is to see whether one arrives at the same outcome using the same parameters as in the original analysis, and the goal of robustness checks is to see whether one arrives at the same outcome using slightly different parameters than in the original analysis. Some authors (Moon et al., 2023) also mentioned that access to data and code is necessary for third parties to redo the analysis on a different dataset.

##### Recommendations for Researchers

Regarding code sharing, recommendations include a modular programming approach, where code is separated into independent and interchangeable modules, version control systems, and a standardization of common analytical approaches (Denaxas et al., 2017). Herrett et al. (2015) point to code repositories for electronic health record research. In these repositories, users can share their methods (metadata, code) so that others can use it or modify it. An example of such a repository is the HDR UK Phenotype Library (https://phenotypes.healthdatagateway.org/).

##### Recommendations for Institutions

Then et al. (2021) would like to see that the conditions of data sharing are stated more clearly when data users make agreements with data providers. Kent et al. (2021) discussed a broader conception of data sharing and argue that data and code availability would not only be useful for other researchers but also for HTA bodies or independent review groups.

##### Limitations

Most discussions about re-sharing data came with caveats, which could be technical, practical, or moral. Many authors cautioned that data sharing is not always allowed because of privacy reasons embedded in data transfer agreements (e.g., Wang et al., 2017; Wang et al., 2019). Many add that information like codebooks or verbal descriptions of the data is necessary for other researchers to effectively re-use the data (e.g., Wang et al., 2017; Wang et al., 2019). To alleviate the concerns surrounding privacy and data transfer agreements, one paper (Matandika et al., 2020) argues in favor of broad consent, where patients provide their consent not only for the original study but also for studies after that, or dynamic consent, where patients can make granular decisions about who can use their data and when.

### Guidance in institutional documents

In total, our sample of 21 institutional documents was slightly different from our preregistered sample, see https://osf.io/m4ehx. We extracted 130 text excerpts from these 21 documents. The prevalence of calls, justifications, and recommendations in those 130 text excerpts is provided in the sections below.

#### Registration

Of the 21 institutional documents, two included a call for more or better registration of SU/HD studies, one included a justification for more or better registration, and seven included one or more recommendations. In total, we found 55 recommendations for the registration of SU/HD studies. The registration recommendations were mainly found in three papers: seven in German Society for Epidemiolog (2008); fifteen in Health Canada (2019); and 24 in Japanese Pharmaceuticals and Medical Devices Agency (2014). Because we found hardly any justifications, recommendations for institutions, and limitations in our set of institutional documents, we only discuss recommendations below.

##### Recommendations for Researchers

The Japanese Pharmaceuticals and Medical Devices Agency (2014) established a ‘Committee for preparation of guidelines on conducting pharmacoepidemiological studies’ to provide a list of elements that would be good to include in registrations of SU/HD studies. Similarly, the German Society for Epidemiology (2008) based their list of to-register elements on a working group, the AGENS Working Group for the Survey and Utilization of Secondary Data, which in turn was strongly based on the Good Epidemiological Practice report, which has been available since 2000 and has seen many revisions, the most recent one in 2019 (Hoffmann et al., 2019). Other documents typically refer to external sources when making recommendations. For example, Health Canada (2019) stated a list of elements that researchers would do well to register based on European Network of Centres for Pharmacoepidemiology and Pharmacovigilance (2010) and International Society of Pharmacoepidemiology (2016), and the Council for International Organizations of Medical Sciences (2024) refers to Berger et al (2007). The specific elements recommended to be included in registrations overlapped greatly with those specified in the peer-reviewed literature.

#### Reporting

For reporting, we found one call and one justification for methods reporting, and one justification for results reporting. We found nineteen recommendations for methods reporting in seven documents, and just one recommendation for results reporting. We only discuss the recommendations in more detail below.

##### Recommendations for Researchers

The Japanese Pharmaceuticals and Medical Devices Agency (2014) and the German Society for Epidemiology (2008) provided several recommendations for methods reporting, most of which revolve around specifying the data source and summarizing the study design. We did not locate recommendations for results reporting aside from a listing of possible publication sites (Council for International Organizations of Medical Sciences, 2024).

#### Sharing

Guidance for data sharing and code sharing was scarce, with only one call for data sharing, two calls for code sharing, and one recommendation for both. We only discuss the recommendations in more detail below.

##### Recommendations for Researchers

While the Council for International Organizations of Medical Sciences (2024) did call for more code sharing and Health Canada (2019) called for more data sharing, they did not provide any recommendations on how to effectively engage in these practices. Only the German Society for Epidemiology (2008) came with some advice: data sharing should only take place with permission of the data owner, but code sharing could take place independently.

## Conclusions and Discussion

In this study, we found that 60 (out of 116) peer-reviewed papers and 12 (out of 23) institutional documents discussed transparency in studies that make secondary use of health data (SU/HD). The justifications for increased transparency included the prevention of questionable research practices, the facilitation of more informative conclusions, and the enhancement of reproducibility and replicability. Recommendations for increased transparency were primarily presented in structured documents like registration templates and reporting guidelines. These documents provide guidance on the study elements to describe in registrations and research papers. For registrations, guidance documents mainly recommended to provide detailed information about the data source, the planned statistical analyses, and any prior knowledge about the data that authors may have. For methods reporting, guidance documents primarily recommended to provide detailed descriptions of the study design, data sources, the variables used in the analysis, and the statistical analysis itself. Recommendations on results reporting highlighted the importance of presenting all conducted analyses, including non-significant results, providing detailed sample characteristics, and running sensitivity analyses. Guidance was limited in the context of data and code sharing. Instead, practical and privacy concerns that could prevent sharing were noted frequently.

Both in the context of registration and reporting, we often found that the data source plays a vital role in the transparency of SU/HD studies. Indeed, some authors argued that existing data, being more voluminous and accessible, provides more scope for researcher biases to creep in, and therefore a higher need for transparency. We agree with that and argue that in the discussions about the transparency of SU/HD studies we should not limit ourselves to methods reporting and results reporting like is typically done for clinical trials but should also include dataset reporting (i.e., providing detailed information about the data source, and the variables in the dataset) as an important pillar. Formalizing these three separate categories of reporting could help researchers become more aware of the need to be transparent in all three of these areas.

As data and code sharing are crucial for reproducibility and transparency, the limited guidance in these areas represents a significant gap that needs to be addressed. In case of data sharing, researchers may not know how to be transparent because the data owner may have placed restrictions on sharing, or it is unclear who is allowed to share and what is allowed to be shared. Further research should prioritize developing detailed and actionable guidelines for data and code sharing in SU/HD studies and how to align these steps with current consent processes.

When recommendations were provided for improving transparency, we often noted that little attention was given to the caveats or concerns related to these recommendations. This is surprising because improving the transparency of a research study is not as straightforward as it may seem. For example, drafting a transparent manuscript typically requires more time and effort than drafting a non-transparent one (Sarafoglou et al., 2022; Spitzer & Mueller, 2023; Tenopir et al., 2011). This can impact researchers at any career stage but is particularly relevant for early career researchers, who rely heavily on producing output quickly to attain their desired academic careers (Allen & Mehler, 2019). Future guidance on improving the transparency of SU/HD studies could discuss the benefits and costs of transparency. When researchers are more aware of the complexities of transparent practices upfront, they may be more likely to continue to engage with transparency practices in the future. If they are faced with challenges during or after improving the transparency of their papers, they may become disgruntled and steer clear of these practices from then on. Research into the day-to-day work processes of researchers may shed more light on this.

Finally, the majority of transparency guidance we found came from papers in the scientific literature. Guidance in institutional documents was relatively sparse, which is important to know because researchers may be more likely to turn to organizations like the WHO or the EMA for guidance than to the peer-reviewed literature. More institutional guidance would align with broader trends in biomedical research that underscore the importance of clear, reproducible, and robust study methodologies to maintain public trust and scientific integrity. In March 2024, a new law came into force in Germany, the Health Data Utilization Act (Gesundheitsdatennutzungsgesetz, GDNG), which regulates the secondary use of health data and, in a separate paragraph, makes both the registration of the corresponding studies in WHO-recognized registers and results reporting mandatory (Bundesministerium für Gesundheit, 2024). One month later, the European Parliament adopted the provisional agreement on the European Health Data Space (EHDS) Regulation (Council of the European Union, 2024). The EHDS will provide researchers, innovators and industry with access to a large health dataset. These developments show that the secondary use of health data is becoming more and more embedded into the scientific ecosystem, highlighting the importance of guidance.

## Limitations

Our review of institutional documents was less comprehensive compared to that of the peer-reviewed literature. While we aimed to include a representative sample of institutional guidance documents, the subjective selection process based on our own expertise may have inadvertently overlooked key documents from health organizations, regulatory bodies, or other relevant entities. This limitation means that our findings might not fully capture the institutional perspective on transparency practices, potentially overlooking valuable insights and recommendations that could have influenced our conclusions.

Relatedly, one could argue that our distinction between peer-reviewed literature and institutional documents was somewhat arbitrary, potentially leading us to underestimate the availability of guidance in institutional documents. Indeed, some peer-reviewed papers involved initiatives or collaborations of formal organizations like the International Society for Pharmacoeconomics and Outcomes Research, ISPOR (Berger et al., 2017; Garrison et al. 2007; Wang et al., 2017). That being said, drawing a line between (more top-down) institutional initiatives and (more bottom-up) initiatives by researchers is hard as it is often difficult to assess the formality of scientific collaborations (e.g. in case of the RECORD initiative, Benchimol et al., 2015; the RECORD-PE initiative, Langan et al., 2018, and the STaRT-RWE initiative, Wang et al., 2021). To allow readers to draw their own conclusions about our sample selection for both peer-reviewed literature and institutional documents, a full list of documents in our sample can be found at https://osf.io/ednwx (peer-reviewed papers) and https://osf.io/gajxt (institutional documents).

Another point to take into consideration when interpreting our results is the subjectivity of the decisions we made throughout the research process. One example lies in our choice of inclusion criteria for guidance documents. While carefully chosen, we might have missed important papers that use different terminology or focus on specific aspects of transparency not covered by our search strategy. Another example of subjectivity in our research choices lies in the nature of our thematic analysis and coding process. Because this process is inherently subjective, our prior knowledge and experience could have introduced bias in our judgments. Although we used established qualitative analysis methods to mitigate this risk, it remains a potential limitation that could affect the reliability of our results. All our quantitative and qualitative codes can be found on the OSF repository of this project: https://osf.io/2nup4.

## General conclusion

Our study highlights substantial efforts in the academic community to enhance transparency in SU/HD studies. To bridge the gap between peer-reviewed recommendations and institutional practices, health organizations could integrate the existing bottom-up initiatives into their formal guidelines. Future research could focus on developing standardized, enforceable guidelines for data and code sharing, while addressing practical and privacy concerns. Additionally, meta-research evaluating the implementation of transparency practices in SU/HD and the impact of transparency practices on research quality, health outcomes and public trust would be desirable.

## Data Availability

All our data is available on the OSF repository of our project at https://osf.io/2nup4. The preregistration is available at https://osf.io/7864h. A document with deviations from our preregistration is available at https://osf.io/m4ehx.

## Author contributions

Conceptualization: OvdA & DS; Data curation: OvdA; Formal analysis: OvdA; Funding acquisition: DS; Investigation: OvdA; Methodology: OvdA, RT, JI, SGS, & DS; Project administration: OvdA; Supervision: DS; Writing – original draft: OvdA; Writing – review & editing: RT, JI, SGS, & DS.

